# Gross motor skills performance in Italian children with and without visual impairment: assess to improve

**DOI:** 10.1101/2025.02.27.25321817

**Authors:** Giulia Chiara Castiglioni, Giulia Hirn, Marco Lippolis, Matteo Porro

## Abstract

The aim of this study is to assess gross motor skills performance of Italian children with and without visual impairment (VI) to quantify the existing gap and to provide an empirical basis to build for VI children a more achieving setting of development to increase their skills for inclusion.

19 VI children (M age = 9,3 years) and 19 sighted children (M age= 8.2 Years) participated. TGMD-2 was used to assess the gross motor skills. The results showed that children with VI had significantly lower both locomotor skills and object control skills than sighted children. Differences were found in relation to the degree of visual impairment: blind children had significantly lower locomotor and object control skills than severe VI children. Practical implications are provided.

WHAT THIS PAPER ADDS? This paper, assessing Italian VI children gross motor skills, collects data useful to make comparisons with results of studies made in other countries and to provide an empirical base for improvement of VI children early intervention programs.

## 1. Introduction

Sight is the primary channel for interacting with the environment and guides and regulates the acquisition, differentiation, and automatization of motor skills. Visual information plays a significant role in motivating children to move, offering details about the distance and direction of movements and objects, assessing potential hazardous situations, and calibrating movement. Additionally, looking at others pushes children to learn movements by example [1].

Impaired visual function during early development significantly influences the progression of motor abilities and the acquisition of skills. From early infancy, children with visual impairment may experience delays in achieving important motor milestones such as controlling their head movements [2]. Tasks like sitting, crawling, standing, and walking are also typically achieved later compared to children with normal vision [3, 4, 5, 6].

Moreover, as children mature, they may encounter challenges related to spatial orientation, temporal coordination, perceiving sensory information, developing body awareness, acquiring self-care skills and making postural adjustment [7, 8, 9, 10].

Visual impairment also negatively impacts gross motor skills, but the studies which analyze this topic are still limited in number, also due to lack of tools validated for visual impaired (VI) children [7]. Gross motor competence is defined as proficiency in a range of fundamental movement skills (e.g. running, throwing, catching, jumping) that are ideally acquired during the preschool and primary school years [11] and plays a significant role in development and providing opportunities for a healthy and active life [12, 13, 14, 15]. Bakke et al. [7] concluded that the only tests that presented validity and reliability of data in studying motor skills of children with visual impairment were Test of Gross Motor Development - 2 (which tests gross motor abilities) and Movement ABC - 2 (which studies manual dexterity, balance and object control). Thus the most suitable test for studying VI children gross motor skills is currently TGMD-2 (available also in a third version not evaluated by Bakke [16]).

Assessing gross motor skills in children is important because subpar motor performance can lead to long term consequences [17, 18]. As Winnick reported [19], adolescents with VI still can have difficulty in performing activities that involve gross motor skills. Poor gross motor performance could have implication in social participation of VI children, considering that gross motor skills serve as the foundational elements for acquiring more intricate movements essential for engaging in sports, games, and actively participating in physical activities [20]. Some studies revealed that targeted activity programs can optimize motor performance in children with VI [4, 21, 22, 23, 24, 25]. Thus, gaining a comprehensive understanding of the qualitative aspects of motor skill performance in VI children is important as it yields valuable insights to design early intervention strategies aimed to avoid enduring ramification of poor motor skills in VI children.

Considering that studies [23, 26, 27, 28] have suggested the existence of variables linked to impaired vision, such as environmental opportunities and barriers for movement, that rather than the visual impairment itself have an impact on the motor skill performance of individuals with VI, we thought that assessing gross motor skill performance in school-aged VI Italian children could add interesting information to the data currently available, mostly derived from foreign countries [23, 29, 30, 31, 32] and therefore from different socio-cultural contexts. At our knowledge there are no previous similar studies made in our country. Italian VI children, since the diagnosis of the visual impairment, are usually included in early rehabilitation programs. Special schools for the blind no longer exist, children are integrated into regular classrooms with their sighted peers. Physical education at school is usually limited at 2 hours per week. Extracurricular sports activities available to VI children are still limited. Inspired by the increasing number of studies about gross motor skills performance in VI children versus sighted children made in other countries, we designed the present study to assess Italian VI children. Main focus is quantifying the existing gap in gross motor skills performance in VI children compared to sighted peers. Data collected can be useful to make comparisons with results of studies made in other countries and to provide an empirical base for improvement of Italian VI children early intervention programs.

## 2. Material and methods

### 2.1 Participants

A total of 38 Italian children were recruited.19 children had visual impairments (age 5 to 12 years, mean age 9.3 years, mean age 111,11 months). 14 were boys and 5 girls. They were recruited from a summer camp organized in Italy by a non-profit Italian sport association (Real Eyes Sport ASD). Only children without other impairments (except from the visual impairment) were selected, based on children’s medical records.

According to the definition of the World Health Organization, based on The International Classification of Diseases 11 (2018), the children were classified as having VI as follow: ten children had a Severe VI (visual acuity worse than 20/200 but equal or better than 20/400). Nine children had Blindness (visual acuity worse than 20/400). 18 of them had a congenital disease and were visual impaired from the birth. 1 of them became blind due to cerebral tumor.

19 sighted children, with an age appropriate to their grade level, were recruited from an Italian leisure center (age 5 to 11 years, mean age 8.2 Years; mean age 98,53 months). 13 were boys, 6 were girls.

### 2.2 Ethical Statement

This study did not require formal ethical approval, as it did not involve any pharmacological or physical interventions, but solely the administration of a standardized motor assessment test.

According to the Comitato Etico dell’Università degli Studi di Milano (Ethics Committee of the University of Milan), ethical approval is not required for observational studies that do not involve interventions or the collection of sensitive data.

All data were fully anonymized, and no participant identifiers were accessible outside the research group, including to hospital staff or the participants themselves.

Informed consent was obtained from the parents or legal guardians of all participants prior to data collection.

The study was conducted in compliance with the ethical principles outlined in the Declaration of Helsinki.

### 2.3 Instruments

#### Test of Gross Motor Development-Second Edition (TGMD-2)

We chose the TGMD-2 to test gross motor performance of participants. TGMD-2 is an instrument widely adopted to test primary-school-age children [33], Houwen et al. [34] also concluded that it’s an appropriate tool to assess the gross motor performance of primary-school-age VI children. It’s validated for children ages 3-10, but already used also for children older than 10 [35].

We adopted the second version of the test (TGMD-2) and not the more recent version TGMD-3 [36] because when the children evaluation was conducted (2020) the TGMD-2 was the one adopted in most studies, especially the ones which comprehended VI children. This could guarantee an easier comparison with results obtained in other countries.

TGMD-2 consists in two subtests that assess locomotor and object control abilities. Each subtest measures 6 gross motor skills. Locomotor subtest assesses skills involved in moving the center of gravity from one point to another: run, gallop, hop, leap, horizontal jump and slide. Object control subtest measures skills involved in throwing and receiving objects: striking a stationary ball, stationary dribbling, catch, kick, overhead throw, underhand roll. Two trials are performed for each gross motor skills and both contribute to the finale score. The examiner measures the quality of the movement using the performance criteria provided by the test manual that assign 0 point if the criterion is absent, 1 point if present. For example, for the Run skills 4 criteria have to be evaluated for each trial, for a total maximum score of 8 points: move in opposition to legs with elbow bent; brief period where both feet are off the ground; foot placement landing on heel or toe (not flat footed); nonsupport leg bent approximately 90 degrees.

TGMD-2 manual provides also normative data, obtained assessing gross motor performance of 1208 persons in 10 US states. Conversion tables provided allow to convert raw scores in standard score (that is raw score standardized by age), percentiles and age equivalent. From standard score it’s possible to calculate gross motor quotient and ‘descriptive ratings’. Descriptive ratings rate the performance of the child (standardized by age) as Very Poor, Poor, Below Average, Average, Above average, Superior, Very Superior, where Average is the score considered normal for age. In the present study, we use raw scores to include in the analyses also children older than 10 years old (N= 5), which is outside the range of TGMD-2 normative data. For these participants we couldn’t calculate standard scores, gross motor score and descriptive ratings, but it was possible to convert raw scores in age equivalent. The original test was adapted to enable children with VI to perform the test, using adjustments inspired by Howen [35]: big, bright, colored cones, instead of normal cones were used marking start and end of the lane and indicating child position; the children were allowed to text the items before the test was administered; due to the impossibility to explain visually the exercises to blind children, if necessary the examiner let the child filled the required movement. Due to the presence of blind children in our study, not included in Howen [35], we designed some special adaptations to enable them to perform some exercises: vocal feedback to keep direction during the locomotor exercises was given by the examiner; bright acoustic balls were used for the object control tests.

### 2.4 Procedure

Each child was tested individually by the same examiner. Sighted children were tested in the playground of the local leisure center. VI children were tested in a sport field during the activities of the summer camp they were attending.

### 2.5 Data analyses

All statistics were performed using “IBM SPSS statistic” software (ver. 28). Descriptive statistics of VI children and sighted children were calculated. The two groups were similar for age and sex.

The dependent variables were locomotor and object control subset raw scores and total raw scores on the TGDM-2. Shapiro-Wilk test was performed to evaluate the distribution of these variables resulting in not normal distributed variables. Due to the result of Shapiro-Wilk test and the low number of children with visual impairment, the nonparametric Mann-Whitney U test was used to compare the variables between children with visual impairments and sighted children. An alpha error < 0.05 were defined to consider significant the difference between all groups.

VI children were divided in two subgroups according to the degree of their deficit: blind children and ones with severe VI. Difference in dependent variables was tested between subgroups, children with severe visual impairment and sighted children, using U test. Finally dependent variables for VI and both subgroups, severe VI and blind, were compared to the results reported in Howen et al. [35] by the Wilcoxon test.

## 3. Results

### 3.1 Gross motor skills of VI children compared to sighted children

VI children differ from sighted children in both the subtests (locomotor and object control) and in the total score. Total score of VI children (Mn = 47.63) is significantly lower than sighted children (Mn = 78.42), p < 0.001. Samely VI children scored significantly lower both in the locomotor (Mn = 25.47) and the object control (Mn = 22.16) subtest than sighted children (LM Mn = 42.26; OC Mn = 36.16), LM p < 0.001, OC p < 0.001) *(Table 1)*. The box and block graphics show as median of Total score, Locomotor and Object control for VI children are significantly lower than sighted children *(Figure 1-3)*.

**Table 1.**
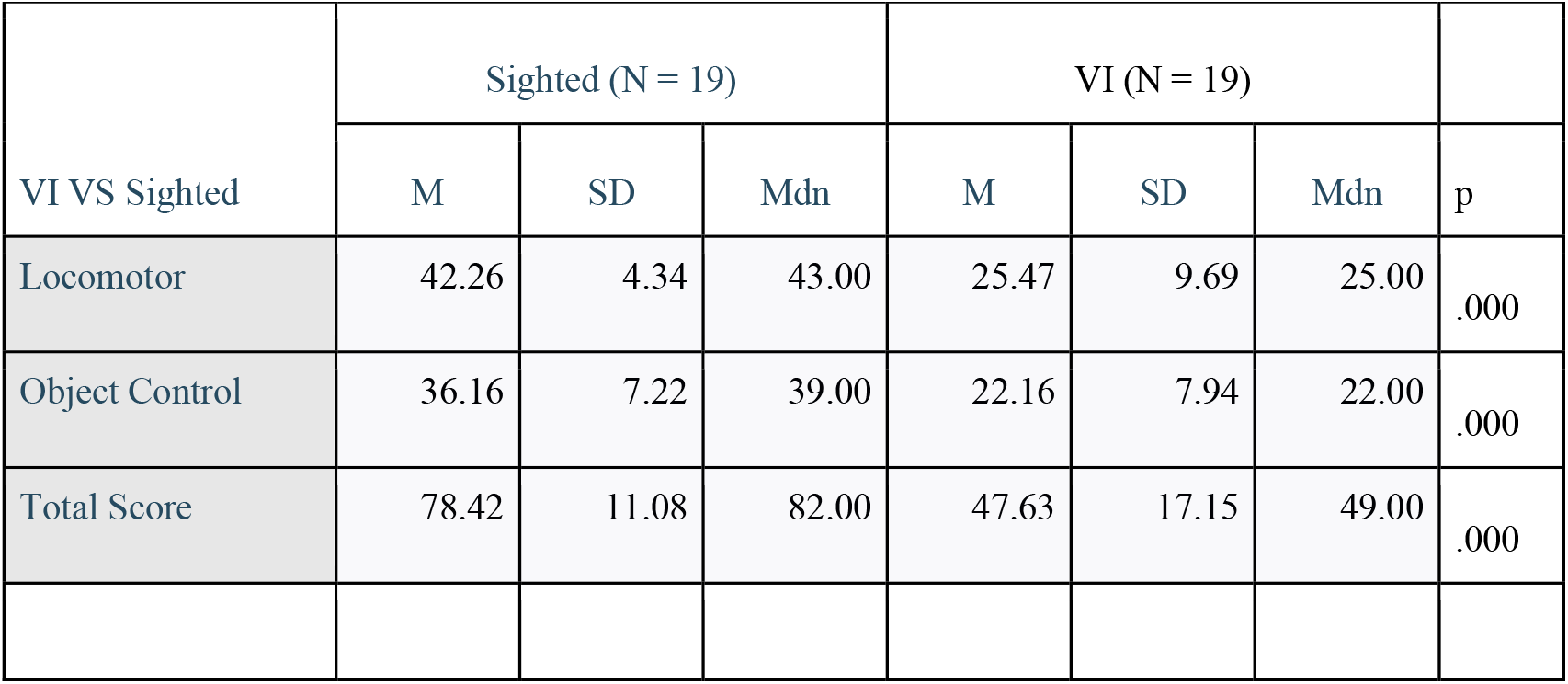
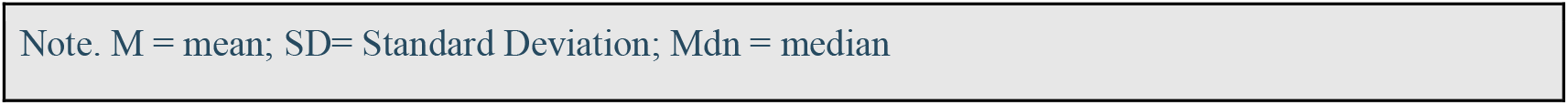
Locomotor and object control scores and total score for VI children and sighted children.

|  |  |  |  |  |  |  |  |
| --- | --- | --- | --- | --- | --- | --- | --- |
| VI VS Sighted | Sighted (N = 19) |  |  | VI (N = 19) |  |  | p |
|  | M | SD | Mdn | M | SD | Mdn |  |
| Locomotor | 42.26 | 4.34 | 43.00 | 25.47 | 9.69 | 25.00 | .000 |
| Object Control | 36.16 | 7.22 | 39.00 | 22.16 | 7.94 | 22.00 | .000 |
| Total Score | 78.42 | 11.08 | 82.00 | 47.63 | 17.15 | 49.00 | .000 |

Note. M = mean; SD= Standard Deviation; Mdn = median

**Figure 1.**
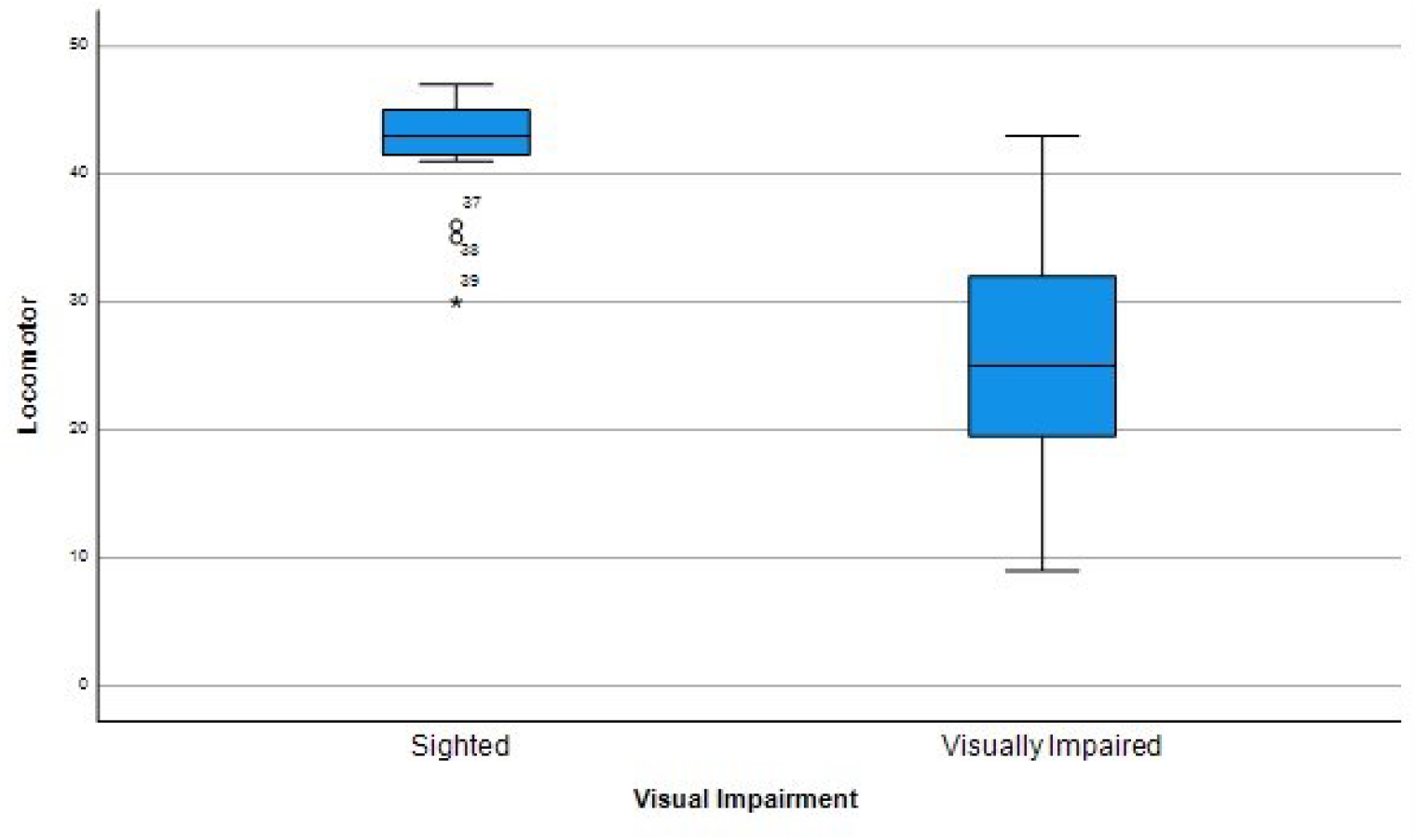

**Figure 2.**
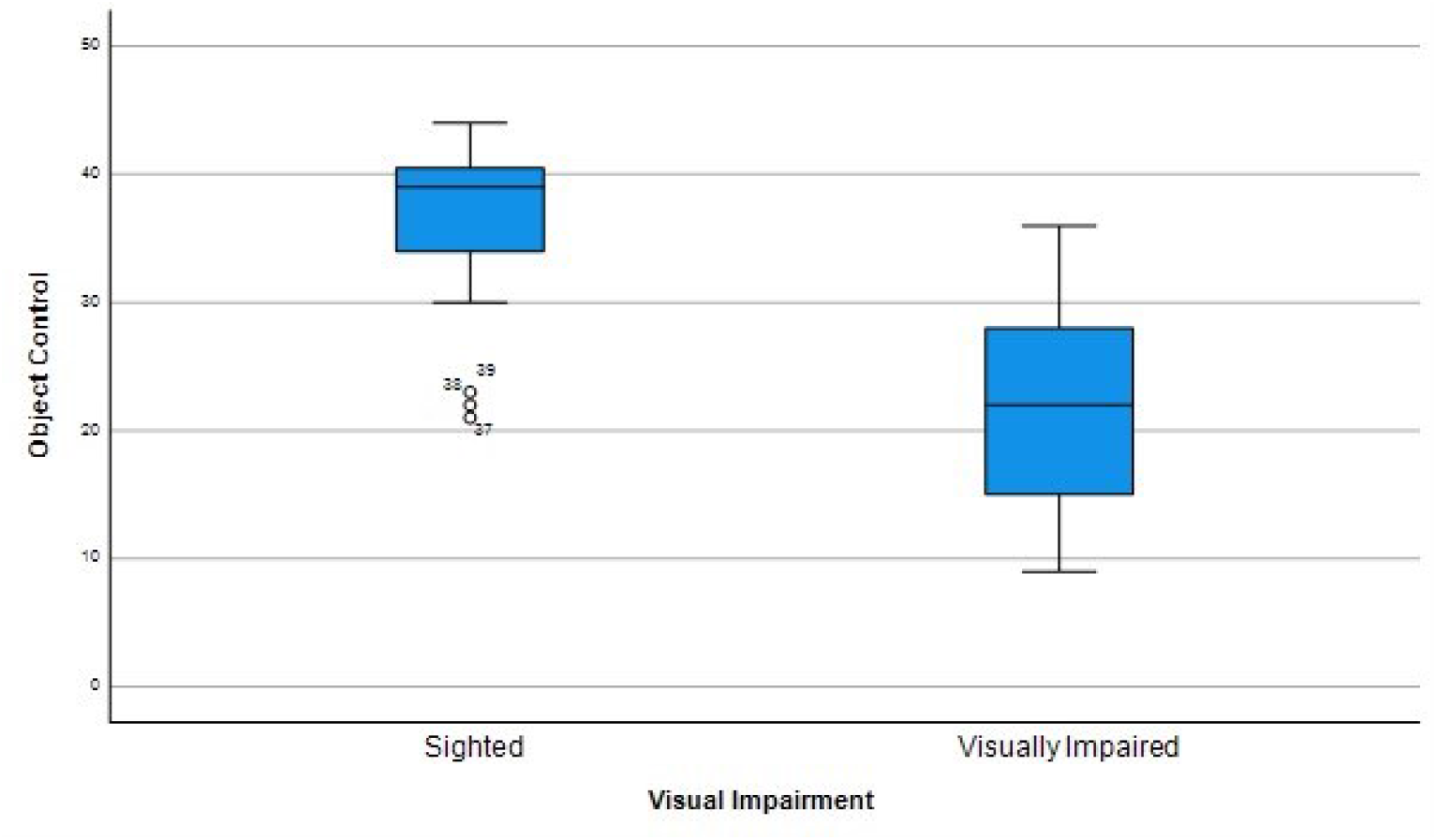

**Figure 3.**
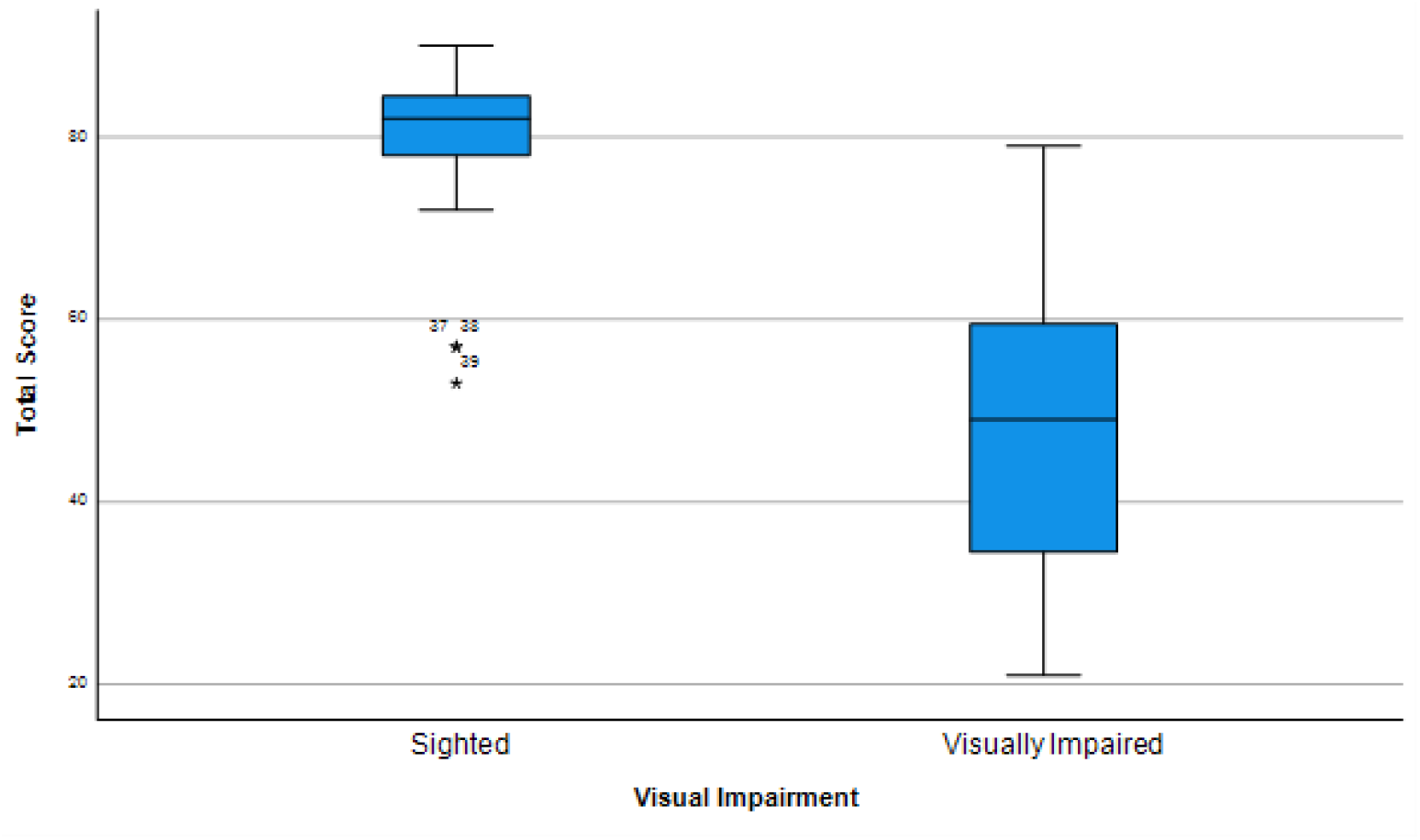

### 3.2 Association of gross motor skills performance and degree of VI children

Since in our study group were recruited children with different degrees of VI, we decided to compare the scores of two subgroups composed by blind children (age 111.0 months, SD 27.16) and severe VI children (age 111.20 months SD 24.9).

Blind children scored significantly lower than severe VI children in both locomotor (p < 0.013) and object control (p < 0.043) subtests and in total score (p < 0.013). *(Table 2)*

**Table 2.** Locomotor and object control scores and total score for children with blindness and with severe VI.

|  |  |  |  |  |  |  |  |
| --- | --- | --- | --- | --- | --- | --- | --- |
|  | Blind (N = 9) |  |  | Severe VI (N = 10) |  |  |  |
|  | M | SD | Mdn | M | SD | Mdn | p |
| Locomotor | 20.00 | 6.16 | 22.00 | 30.40 | 9.84 | 32.00 | .000 |
| Object Control | 18.11 | 5.95 | 17.00 | 25.80 | 7.97 | 28.00 | .000 |
| Total Score | 38.11 | 11.57 | 37.00 | 56.20 | 17.24 | 59.5 | .000 |
| Note. Md = mean; SD= Standard Deviation; Mdn = median |  |  |  |  |  |  |  |

### 3.3 Gross motor skills of children with severe VI compared to sighted children

The observation of a great difference in gross motor performance between blind children group and severe VI ones shown in Table 2, suggested us to compare the differences between the scores of sighted children and severe visual impaired ones.

Severe VI children scores are significantly lower than sighted children in locomotor (p < 0.001), object control (p = 0.001) and total score test (p < 0.001). *(Table 3)*

**Table 3.** Locomotor and object control scores and total score for severe VI children and sighted children.

|  |  |  |  |  |  |  |  |
| --- | --- | --- | --- | --- | --- | --- | --- |
|  | Sighted (N = 19) |  |  | Severe VI (N = 10) |  |  | p |
|  | M | SD | Mdn | M | SD | Mdn |  |
| Locomotor | 42.26 | 4.34 | 43.00 | 30.40 | 9.84 | 32,00 | .000 |
| Object Control | 36.16 | 7.22 | 39.00 | 25.80 | 7.97 | 28.00 | .000 |
| Total Score | 78.42 | 11.08 | 82,00 | 56.20 | 17.24 | 59.5 | .000 |
| Note. M = mean; SD= Standard Deviation; Mdn = median |  |  |  |  |  |  |  |

### 3.4 Gross motor skills of Severe VI Italian children compared to Severe VI Dutch children

We deemed it interesting to compare the scores obtained by the participants of our study with ones assessed in other countries. Among the few studies that used TGMD-2 to analyze gross motor skills of VI children, we considered Howen et al. [35] the most suitable for a comparison, because it reports Mean, Median and SD of the subtest and total raw score of VI Dutch children in a similar age range (6-11, mean age 9.20 years) than our participants. Moreover, we used similar adaptations of TGMD-2 to enable VI children to perform TGMD-2 trials. In Howen et al. [35] the sample tested can be divided into two subgroups: children with moderate VI (N= 7) and children with severe VI (N=7) according to WHO definition; there were no children with blindness tested. We decided then to compare our participants with Severe VI with Severe VI Dutch children.

Italian children scores significantly lower than Dutch children in locomotor subtest (p = 0.015) and in total score (p = 0.02), while they don’t seem to differ from Dutch children in OC score (p = 0.10). (Table 4)

**Table 4.** Locomotor and object control scores and total score for Italian Severe VI children and Dutch Severe VI children.

|  |  |  |  |  |  |  |  |
| --- | --- | --- | --- | --- | --- | --- | --- |
|  | Italian Severe VI (N = 10) |  |  | Dutch Severe VI (N = 7) |  |  |  |
|  | M | SD | Mdn | M | SD | Mdn | p |
| Locomotor | 30.40 | 9.84 | 32.00 | 39.3 | 2.6 | 39.0 | 0.015 |
| Object Control | 25.80 | 7.97 | 28.00 | 32.7 | 5.9 | 31.0 | 0.10 |
| Total Score | 56.20 | 17.24 | 59.5 | 72.0 | 7.7 | 71.0 | 0.02 |
| Note. M = mean; SD= Standard Deviation; Mdn = median |  |  |  |  |  |  |  |

### 3.5 Gross motor skills performance of Italian VI children compared with TGMD-2 normative data

Table 5 show “age equivalent”, “gross motor score” and “descriptive ratings” which describe the global gross motor performance of each VI children aged 3 to 10 years. Raw scores and descriptive ratings of each subtest (locomotor and object control) are reported too. For five participants was not possible to calculate “gross motor quotient” and “descriptive ratings”, because they exceed age limit of 10 year and 11 months. Instead, it was possible (and indeed it was considered appropriate and interesting) to indicate for these subjects the age equivalent based on the raw scores obtained. As shown in Graph 3, 9 children (corresponding to 47% of VI children) obtained a descriptive rating of “Very Poor” in the global score, 4 children (21%) scored “Poor”, 1 child (5%) obtained a “Below average” descriptive rating. Graph 1 and Graph 2 show the descriptive ratings of each subtest (locomotor and object control). Very Poor corresponds to a Gross Motor Quotient of less than 70, which is a value found only in 2.34% of the normative population without visual disabilities. The average calculated Gross Motor Quotient is approximately 61.5, equivalent to a percentile below 1 in the normative American population without visual impairment.

**Table 5.**

| Locomotor subtest |  |  |  |  | Object control subtest |  |  | Total score |  |
| --- | --- | --- | --- | --- | --- | --- | --- | --- | --- |
| Sub<br>ject | Age | Raw<br>score | Age<br>equivalent | Descriptive<br>ratings | Raw<br>score | Age<br>equivalent | Descriptive<br>ratings | QGM | Descriptive<br>ratings. |
| 1 | 12y<br>0m-12 y<br>11m | 30 | 5 y - 0<br>m | / | 32 | 6 y 6<br>m | / | / | / |
| 2 | 11y<br>0m-11 y<br>11m | 39 | 6y -<br>6m | / | 34 | 5 y 9 m | / | / | / |
| 3 | 11y<br>0m-11y<br>11m | 13 | <3 y | / | 12 | <3 y | / | / | / |
| 4 | 10y<br>0m-10y<br>11m | 28 | 4 y -<br>6m | Very poor | 25 | 5 y | Very poor | 58 | Very poor |
| 5 | 11y<br>0m-11y<br>11m | 23 | 3 y -<br>9m | / | 26 | 4y 6 m | / | / | / |
| 6 | 11y<br>0m-11y<br>11m | 25 | 4 y - 0<br>m | / | 22 | 4 y 3 m | / | / | / |
| 7 | 10y<br>0m-10y<br>11m | 43 | 8 y - 6<br>m | Average | 36 | 7 y 6 m | Poor | 82 | Below<br>average |
| 8 | 9y<br>0m-9y<br>11m | 9 | < 3 y | Very poor | 14 | < 3 y | Very poor | 46 | Very poor |
| 9 | 9y<br>0m -<br>9y<br>11m | 34 | 5 y 6<br>m | Very poor | 30 | 5 y 6 m | Poor | 64 | Very poor |
| 10 | 9y<br>0m-9y<br>11m | 36 | 6 y 0<br>m | Below<br>average | 30 | 6 y 0 m | Below<br>average | 76 | Poor |
| 11 | 9y<br>0m -<br>9y<br>11m | 33 | 5 y 6<br>m | Poor | 22 | 4 y 3 m | Very poor | 61 | Very poor |
| 12 | 9y<br>0m -<br>9y<br>11m | 20 | 3 y 3<br>m | Very poor | 17 | < 3 y | Very poor | 46 | Very poor |
| 13 | 8y<br>0m –<br>8y<br>11m | 31 | 5 y 0<br>m | Poor | 26 | 4 y 6 m | Very poor | 61 | Very poor |
| 14 | 7y<br>0m –<br>7y<br>11m | 25 | 4 y 0<br>m | Very poor | 22 | 3 y 9 m | Very poor | 55 | Very poor |
| 15 | 8y<br>0m –<br>8y<br>11m | 12 | <3 y | Very poor | 9 | <3 y | Very poor | 46 | Very poor |
| 16 | 7y<br>0m –<br>7y 11<br>m | 12 | <3 y | Very poor | 16 | <3 y | Very poor | 46 | Very poor |
| 17 | 6y<br>0m –<br>6y<br>11m | 30 | 5 y 0<br>m | Below<br>average | 20 | 4 y 0 m | Poor | 76 | Poor |
| 18 | 5y<br>0m-5y<br>11m | 19 | 3 y 0<br>m | Below<br>average | 14 | <3 y | Poor | 70 | Poor |
| 19 | 5y<br>0m –<br>5y<br>11m | 22 | 3 y 6<br>m | Below<br>average | 14 | <3 y | Poor | 73 | Poor |

Rounding up to 3 years and 0 months the performance of children who obtained an age equivalent score of <3 years, we calculated that the estimated age of VI children based on gross motor performance is on average lower than their chronological age (and therefore below the normative data for a normally sighted subject) by 58 months (equivalent to 4 years and 10 months) in locomotor skills, and at least 59 months, that is, 4 years and 11 months, in object control skills.

**Graph 1.**
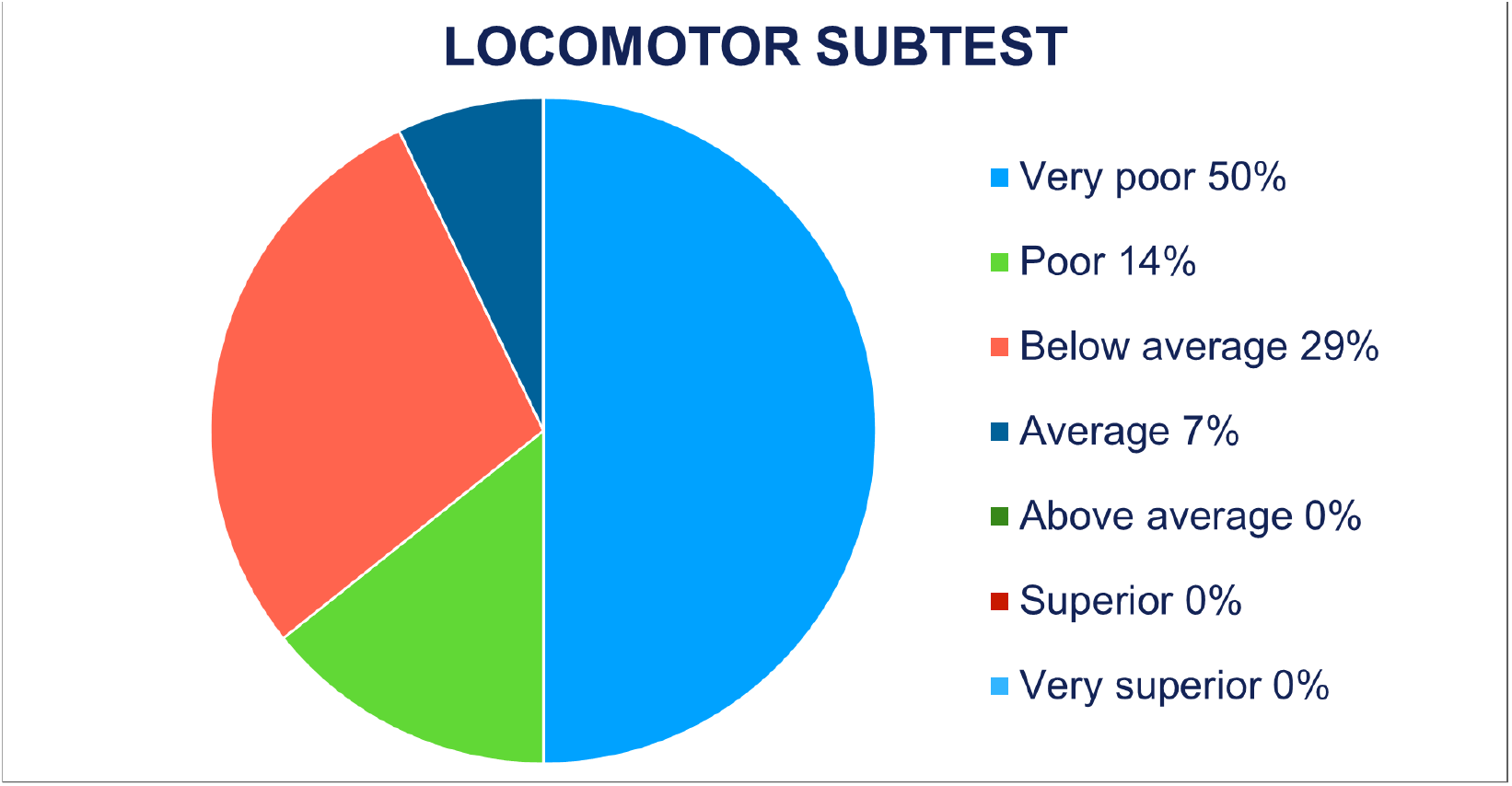

**Graph 2.**
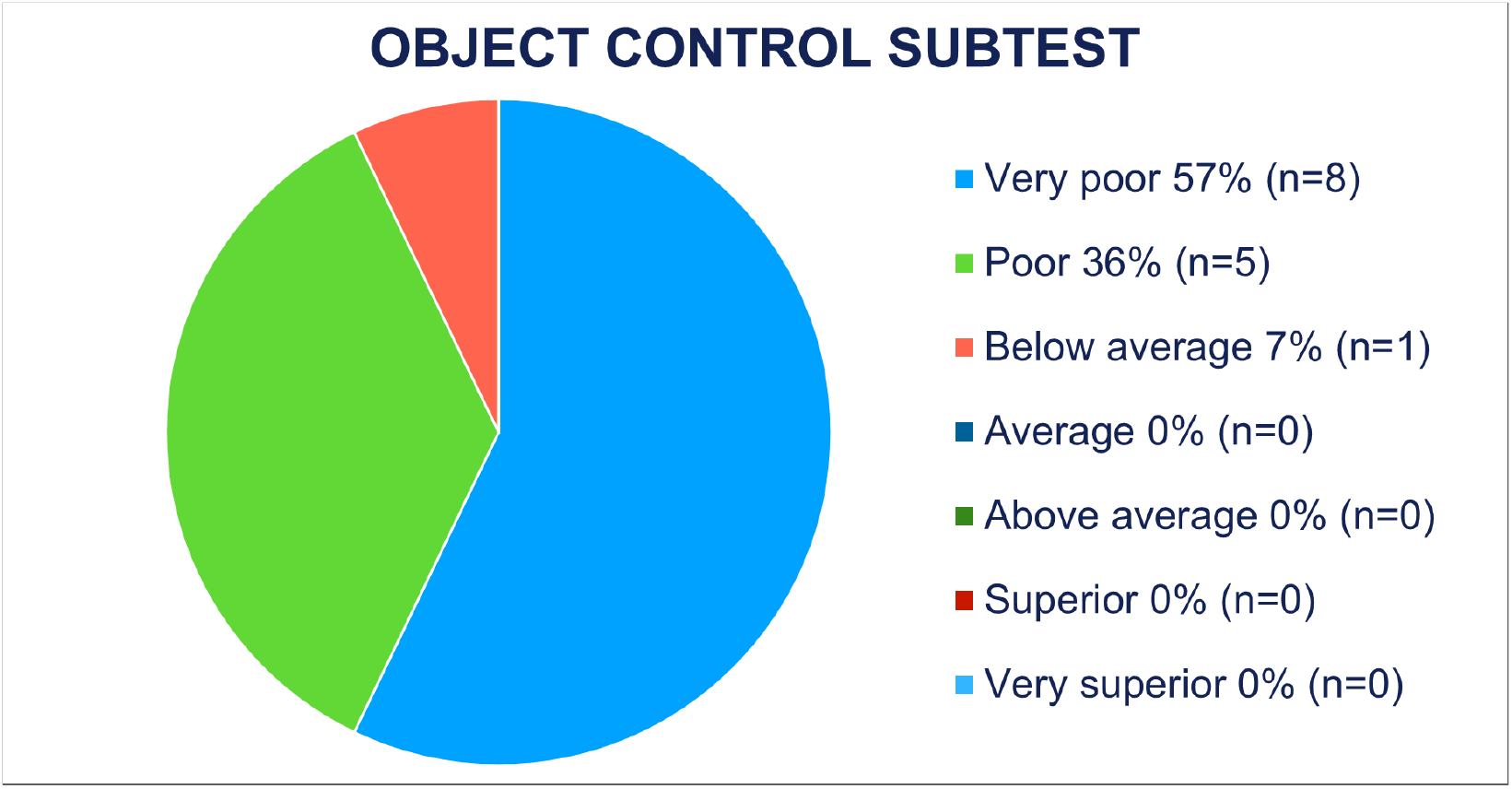

**Graph 3.**
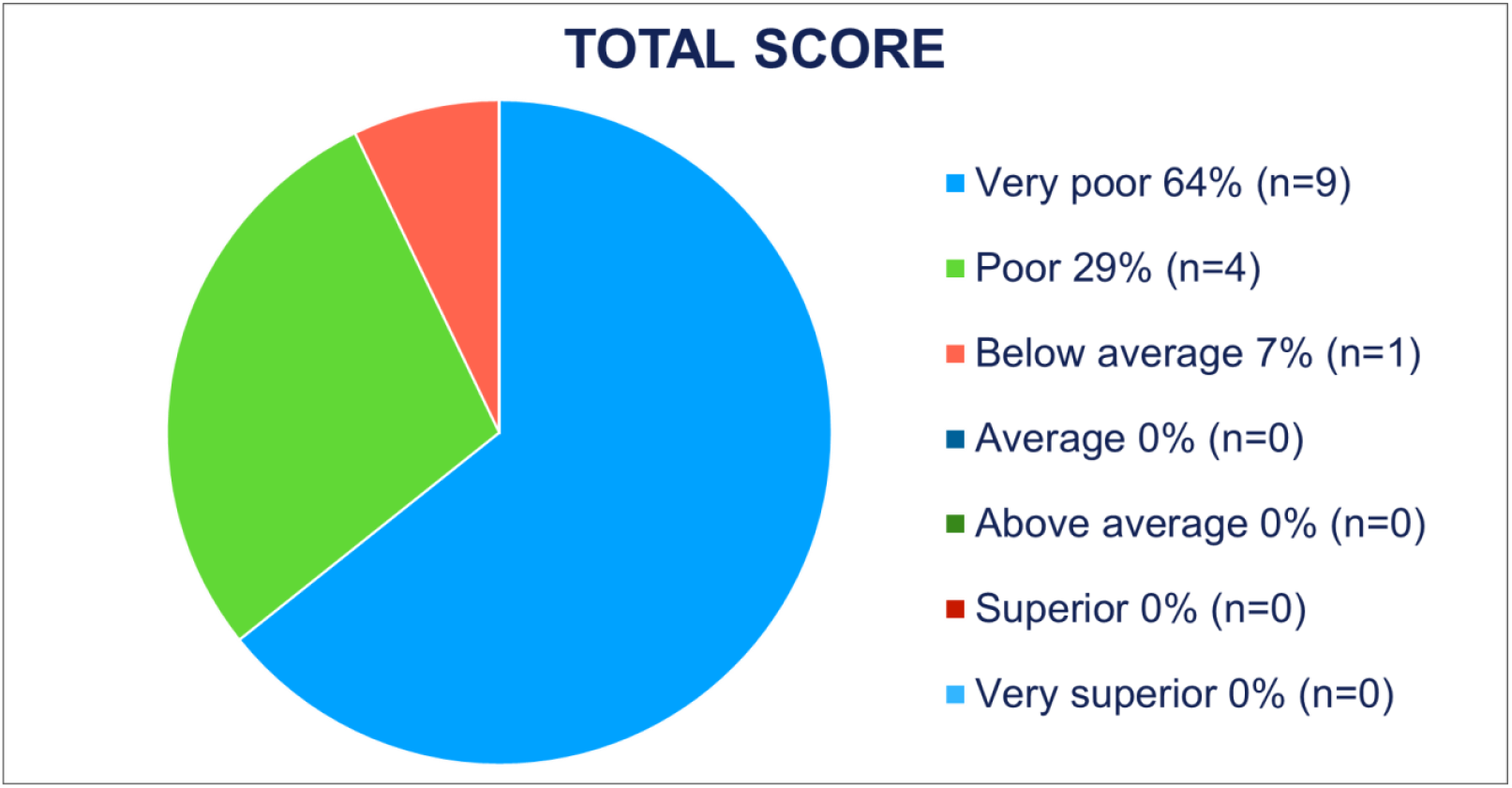

## 4. Discussion

This is the first study to investigate gross motor skills performance in Italian school aged children with and without VI. Our findings suggest that Italian children with VI have significantly lower gross motor skills compared to sighted Italian children of the same age, as well as compared to normative data from the test (reflecting the American norm population without visual impairment [33]. This finding is consistent with previous studies that underline the significant role of the visual channel in gross motor skills acquisition [1, 2, 3, 4, 5, 6]. However, we deemed it necessary to further investigate and quantify the observed gap. Firstly, the difference in motor performance between sighted and VI kids was recorded both in locomotor skills and object control. This latter finding partially deviates from a previous study [35] conducted on Dutch children, which showed differences between sighted and VI children in object control abilities but not in locomotor skills. The authors concluded that the good performance achieved by VI children in the locomotor subtest could be explained by the fact that locomotor skills are often utilized by VI children in their daily motor play and sports activities. The subjects we studied, with comparable visual deficits, exhibit significant difficulties even in the locomotor subtest, suggesting that these individuals do not frequently experience these gestures in their daily lives.

Regarding object control skills, it is plausible that VI children demonstrate lower performance due to the importance of the visual channel in providing feedback on object position and movement [35,19].

It is also true that controlling objects such as balls (using hands as in the throwing task or using feet as in the kicking task) are common and fundamental gestures in the play activities of sighted children. Therefore, it is evident how the lack of mastery of these gestures can be detrimental in terms of inclusion.

The degree of visual acuity was found to influence motor performance in both the locomotor subtest and object control subtest: blind children tested had a lower gross motor skill level than severe VI children, which can be expected given the possibility for the latter to rely partially on the visual channel [35]. However, even with better visual acuity, the children with severe visual impairment were unable to achieve motor performance comparable to that of their sighted peers.

To quantify the developmental gap that emerged in motor skills between VI and sighted children, we utilized the age-equivalent calculation, percentiles, and gross motor quotient provided by the TGMD-2 test. The results are quite alarming: the average level of motor ability recorded in the group of VI children is found in less than 1% of the normative population without visual impairment. The motor gap between the two categories, within the limitations of the instrument, can be quantified as over 4 years on average. Although it is well-established that motor milestones in VI children are achieved later compared to their sighted peers [3, 4, 5, 6], we questioned whether this gap may never be fully closed if it is too pronounced, especially considering that the critical period for the acquisition of gross motor skills occurs during the early years of life [37].

The calculation of age equivalence (table 5) demonstrates that even VI children over the age of 10 show a lower “gross motor age” by several years compared to their sighted peers (on which the normative data is based). It is clear that an 11-year-old child with a gross motor age equivalent to that of a 5-year-old will face significant challenges in participating in motor play activities with peers of the same age. Improving motor skills at the age of 11 will also be more difficult compared to intervening during the early years of life.

Comparing the data obtained with those collected in other countries, such as the study by Howen [35] it appears that there is significant room for improvement for these children: Dutch children with similar visual impairment show significantly higher scores compared to Italian children. Despite the limited number of subjects studied in both studies, it is reasonable to believe that increasing motor activity in VI children during the early years of life can lead to motor skills that are less disparate from their sighted peers, thus promoting inclusion in motor and sports activities. The same Dutch study shows that VI children who participate in organized sports activities demonstrate better object control compared to those who do not participate. Additionally, other studies highlight the benefits of physical activity on gross motor skills in VI children [7, 22, 23, 25, 38, 39].

Warren [26] emphasized the importance of providing VI children with ample opportunities to experience movement. Therefore, we assume that early participation in recreational motor activities and sport activities among Italian VI children could be a path to improve their gross motor skills and reduce the gap compared to their sighted peers. These activities, if appropriately designed and made accessible, could complement the rehabilitative interventions. Sports and recreational motor activities offer the advantage, compared to rehabilitative activities, of allowing children to experience movement in a playful context with their peers, thus being more motivating and fostering social relationships. It is also important to note that unstructured motor activity, such as daily free play at home, in playgrounds, or at preschool, can play an important role in increasing children’s motor activity levels [28].

Educating families to pay more attention to the daily movement levels of their VI children, even in the early years of life, could be key to promote the development of gross motor skills.

## 5. Limitations and future perspectives

The limitations of the study are the small sample size and the lack of an assessment tool specifically designed to test gross motor skills in VI children. In fact, despite the validation of TGMD-2 according to Howen [35], even for VI children, we encountered objective difficulties in administering some tests to blind children, particularly the object control tests. Furthermore, more precise instructions on how to explain the exercises to blind children would be necessary.

It would be interesting in future studies to precisely investigate the levels of daily movement in Italian VI children and assess how much gross motor performance can improve after a period of exposure to appropriately structured motor activities. Additionally, exploring the impact of poor gross motor performance on autonomy and social inclusion would also be of interest.

## Data Availability

All data produced in the present study are available upon reasonable request to the authors

## 6. Acknowledgements

We are grateful to the infants and families that participated in the study. We also thank Real Eyes Sport association for its contributions. In particular, we especially thank Daniele Cassioli, PT, paralympic athlete and president of Real Eyes Sport, for believing in the potential of children, prioritizing their skills over their disabilities.

## References

[1] M. Brambring, “Divergent Development of Gross Motor Skills in Children who are Blind or Sighted,” J Vis Impair Blind, vol. 100, no. 10, pp. 620–634, 2006.

[2] H. F. R. Prechtl, G. Cioni, C. Einspieler, A. F. Bos, and F. Ferrari, “Role of vision on early motor development: lessons from the blind,” Dev Med Child Neurol, vol. 43, no. 3, pp. 198–201, 2001.

[3] A. Hallemans, E. Ortibus, S. Truijen, and F. Meire, “Development of independent locomotion in children with a severe visual impairment,” Res Dev Disabil, vol. 32, no. 6, pp. 2069–2074, 2011.

[4] F. Elisa et al., “Gross motor development and reach on sound as critical tools for the development of the blind child.” [Online]. Available: www.elsevier.com/locate/braindev

[5] O. Levtzion-Korach, A. Tennenbaum, R. Schnitzer, and A. Ornoy, “Early motor development of blind children,” J Paediatr Child Health, vol. 36, no. 3, pp. 226–229, 2000.

[6] E. Adelson and S. Fraiberg, “Gross Motor Development in Infants Blind from Birth,” Child Dev, vol. 45, no. 1, pp. 114–126, 1974.

[7] H. A. Bakke, W. A. Cavalcante, I. S. de Oliveira, S. W. Sarinho, and M. T. Cattuzzo, “Assessment of Motor Skills in Children With Visual Impairment: A Systematic and Integrative Review,” Clin Med Insights Pediatr, vol. 13, p. 117955651983828, Jan. 2019, doi: 10.1177/1179556519838287.

[8] PILAR ARNAIZ SANCHEZ, Deficiencias visuales y psicomotricidad: teoría y práctica, 1st edition. Madrid, 1994.

[9] M. R. Matos, “Análise do equilíbrio em postura ortostática em crianças com deficiência visual por meio de parâmetros estabilométricos,” São José dos Campos, Brazil, 2006.

[10] M. Matos, C. Matos, and C. Oliveira, “Equilíbrio estático da criança com baixa visão por meio de parâmetros estabilométricos,” Fisioterapia em Movimento, vol. 23, pp. 361–369, 2010.

[11] D. R. Lubans, P. J. Morgan, D. P. Cliff, L. M. Barnett, and A. D. Okely, “Fundamental Movement Skills in Children and Adolescents Review of Associated Health Benefits.”

[12] C. Branta, J. Haubenstricker, and V. Seefeldt, “Age Changes in Motor Skills During Childhood and Adolescence,” Exerc Sport Sci Rev, vol. 12, no. 1, pp. 467–520, Jan. 1984, doi: 10.1249/00003677-198401000-00015.

[13] D.L., & O. J. C. Gallahue, Understanding motor development : infants, children, adolescents, adults, 6th ed. Boston, 2006.

[14] D. L. Gallahue and F. C. Donnelly, “Developmental physical education for all children,” Palaestra, vol. 20, no. 1, 2003.

[15] L. M. Barnett et al., “Correlates of Gross Motor Competence in Children and Adolescents: A Systematic Review and Meta-Analysis,” Sports Medicine, vol. 46, no. Springer International Publishing, pp. 1663–1688, Nov. 01, 2016. doi: 10.1007/s40279-016-0495-z.

[16] D. A. Ulrich, “Introduction to the special section: Evaluation of the psychometric properties of the TGMD-3,” J Mot Learn Dev, vol. 5, no. 1, pp. 1–4, Jun. 2017, doi: 10.1123/jmld.2017-0020.

[17] M. H. Cantell, M. M. Smyth, and T. P. Ahonen, “Clumsiness in Adolescence: Educational, Motor, and Social Outcomes of Motor Delay Detected at 5 Years,” Human Kinetics Publishers, Inc, 1994.

[18] M. Cousins and M. M. Smyth, “Developmental coordination impairments in adulthood,” Hum Mov Sci, vol. 22, no. 4–5, pp. 433–459, Nov. 2003, doi: 10.1016/j.humov.2003.09.003.

[19] J. P. Winnick, “The Performance of Visually Impaired Youngsters in Physical Education Activities: implications for Mainstreaming,” 1985.

[20] A. Brian, N. Getchell, L. True, A. De Meester, and D. F. Stodden, “Reconceptualizing and Operationalizing Seefeldt’s Proficiency Barrier: Applications and Future Directions,” Sports Medicine, vol. 50, no. 11. Springer Science and Business Media Deutschland GmbH, pp. 1889–1900, Nov. 01, 2020. doi: 10.1007/s40279-020-01332-6.

[21] E. Aki, S. Atasavun, A. Turan, and H. Kayihan, “Training motor skills of children with low vision.,” Percept Mot Skills, vol. 104, pp. 1328–1336, 2007.

[22] E. Fazzi, S. G. Signorini, S. M. Bova, P. Ondei, and P. E. Bianchi, “Early intervention in visually impaired children,” Int Congr Ser, vol. 1282, pp. 117–121, Sep. 2005, doi: 10.1016/j.ics.2005.05.200.

[23] Houwen Susanne, Visscher Chris, Lemmink KAPM, and Hartman Ester, “Motor skill performance of children and adolescents with visual impairments: a review,” Except Child, vol. 75, pp. 464–492, 2009.

[24] Jazi Shirin Davarpanah, Purrajabi Fatemeh, Movahedi Ahmadreza, and Jalali Shahin, “Effect of selected balance exercises on the dynamic balance of children with visual impairments,” J Vis Impair Blind, vol. 106, pp. 466–474, 2012.

[25] L. Santos, J. Passos, and A. Rezende, “Os efeitos da aprendizagem psicomotora no controle das atividades de locomoção sobre obstáculos em crianças com deficiencia da visão.,” Revista Brasileira de Educação Especial, vol. 13, pp. 365–380, 2007.

[26] D. H. Warren, Blindness and children: An individual differences approach. Cambridge University Press, 1994.

[27] J. C. Ozmun and D. L. Gallahue, Understanding Motor Development: Infants, Children, Adolescents, Adults, 6th ed. New York: McGraw-Hill, 2006.

[28] L. H. Schneekloth, “Play Environments for Visually Impaired Children,” J Vis Impair Blind, vol. 83, no. 4, pp. 196–201, 1989.

[29] P. S. Haibach, M. O. Wagner, and L. J. Lieberman, “Determinants of gross motor skill performance in children with visual impairments,” Res Dev Disabil, vol. 35, no. 10, pp. 2577–2584, 2014, doi: 10.1016/j.ridd.2014.05.030.

[30] S. Houwen, E. Hartman, and C. Visscher, “The Examining relationship among motor proficiency, physical fitness, and body composition in children with and without visual impairments,” Res Q Exerc Sport, vol. 81, pp. 291–300, 2010.

[31] M. O. Wagner, P. S. Haibach, and L. J. Lieberman, “Gross motor skill performance in children with and without visual impairments-Research to practice,” Res Dev Disabil, vol. 34, no. 10, pp. 3246–3252, Oct. 2013, doi: 10.1016/j.ridd.2013.06.030.

[32] G. F. Fard, J. M. Oori, and H. N. Zadeh, “Psychometric Properties of Gross Motor Development Test (Second Edition) in Children with Visual Impairment in Tehran of Test of Gross Motor Development and second edition of Movement Assessment Battery for Children,” Journal of Paramedical Sciences & Rehabilitation, vol. 9, no. 3, pp. 31–38, 2020, [Online]. Available: http://jpsr.mums.ac.ir

[33] D. A. Ulrich, “Test of gross motor development (2nd ed.).” PRO-ED, Austin, 2000.

[34] S. Houwen, E. Hartman, L. Jonker, and C. Visscher, “Reliability and Validity of the TGMD-2 in Primary-School-Age Children With Visual Impairments,” Adapted Physical Activity Quarterly, vol. 27, no. 2, pp. 143–159, 2010.

[35] S. Houwen, C. Visscher, E. Hartman, and Lemmink Koen APM, “Gross Motor Skills and Sports Participation of Children With Visual Impairments,” Res Q Exerc Sport, vol. 78, pp. 16–23, 2007.

[36] D. A. Ulrich, “Introduction to the special section: Evaluation of the psychometric properties of the TGMD-3,” J Mot Learn Dev, vol. 5, no. 1, pp. 1–4, Jun. 2017, doi: 10.1123/jmld.2017-0020.

[37] F. M. Murphy and M. O’Driscoll, “Observations on the Motor Development of Visually Impaired Children Interpretations from Video Recordings,” Physiotherapy (United Kingdom), vol. 75, no. 9, pp. 505–508, 1989, doi: 10.1016/S0031-9406(10)62300-0.

[38] C. Graf, B. Koch, E. Kretsschmann-Kandel, G. Falkowski, H. Christ, and S. Coburger, “Correlation between BMI, leisure habits and motor abilities in childhood (CHILT - Project),” Int J Obes, vol. 28, pp. 22–26, 2004.

[39] A. D. Okely, M. L. Booth, and J. W. Patterson, “Relationship of physical activity to fundamental movement skills among adolescents,” Medicine & Science in Sport & Exercise, vol. 33, pp. 1899–1904, 2001.

